# Aberrant stromal tissue factor and mycolactone-driven vascular permeability, exacerbated by IL-1β, orchestrate pathogenic fibrin formation in Buruli ulcer lesions

**DOI:** 10.1101/2021.08.04.21261598

**Authors:** Louise Tzung-Harn Hsieh, Scott J Dos Santos, Joy Ogbechi, Aloysius D. Loglo, Francisco J. Salguero, Marie-Thérèse Ruf, Gerd Pluschke, Rachel E. Simmonds

## Abstract

The neglected tropical disease Buruli ulcer, caused by *Mycobacterium ulcerans* infection, displays coagulative necrosis in affected skin tissues. We previously demonstrated that exposure to the *M. ulcerans* exotoxin mycolactone depletes the expression of thrombomodulin and impacts anticoagulation at the endothelial cell surface. Moreover, while widespread fibrin deposition is a common feature of BU lesions, the cause of this phenotype is not clear. Here, we performed sequential staining of serial tissue sections of BU patient biopsies and unbiased analysis of up to 908 individual non-necrotic vessels of eight BU lesions to investigate its origins. Most vessels showed evidence of endothelial dysfunction being thrombomodulin-negative, von Willebrand factor-negative and/or had endothelium that stained positively for tissue factor (TF). Primary haemostasis was rarely evident by platelet glycoprotein CD61 staining. Localisation of TF in these lesions was complex and aberrant, including diffuse staining of the stroma some distance from the basement membrane and TF-positive infiltrating cells (likely eosinophils). This pattern of abnormal TF staining was the only phenotype that was significantly associated with fibrin deposition, and its extent correlated significantly with the distance that fibrin deposition extended into the tissue. Hence, fibrin deposition in Buruli ulcer lesions is likely driven by the extrinsic pathway of coagulation. To understand how this could occur, we investigated whether clotting factors necessary for fibrin formation might gain access to the extravascular compartment due to loss of the vascular barrier. *In vitro* assays using primary vascular and lymphatic endothelial cells showed that mycolactone increased the permeability of monolayers to dextran within 24 hours. Moreover, co-incubation of cells with interleukin-1β exacerbated mycolactone’s effects, nearly doubling the permeability of the monolayer compared to each challenge alone. We propose that leaky vascular and lymphatic systems are important drivers of extravascular fibrin deposition, necrosis and oedema frequently seen in Buruli ulcer patients.

**Author Summary:** To date, the debilitating skin disease Buruli ulcer remains a public health concern and financial burden in low or middle-income countries, especially in tropical regions. Late diagnosis is frequent in remote areas, perhaps due to the painlessness of the disease. Hence patients often present with large, destructive opened ulcers leading to delayed wound closure or even lifelong disability. The infectious agent produces a toxin called mycolactone that drives the disease. We previously found evidence that the blood clotting system is disrupted by mycolactone in these lesions, and now we have further explored potential explanations for these findings by looking at the expression of coagulation regulators in BU. In detailed analysis of patient skin punch biopsies, we identified distinct expression patterns of certain proteins and found that tissue factor, which initiates the so-called extrinsic pathway of blood clotting, is particularly important. Mycolactone is able to disrupt the barrier function of the endothelium, further aggravating the diseased phenotype, which explains how clotting factors access the tissue. Altogether, such localised hypercoagulation in Buruli ulcer skin lesions may contribute to the development of the lesion.

## Introduction

Buruli ulcer (BU) is a neglected tropical disease, resulting from a subcutaneous infection with *Mycobacterium ulcerans*, with approximately 2700 new cases reported globally in 2018 according to the latest update from World Health Organization (WHO) [1]. BU endemic areas include West and Central African countries as well as Australia; globally over 30 countries have reported cases with estimated prevalence ranging from 3.2-26.7 BU cases per 10,000 population [2, 3]. BU presents in various forms such as nodules, plaques, ulcers and oedema [2]. In late diagnosed cases, necrotic skin lesions and soft tissue destruction can extend to 15% of body surface area, which may cause lifelong disability [4]. This debilitating skin condition can be treated with dual anti-mycobacterial antibiotics, sometimes alongside surgical removal of infected skin tissues [5]. Yet delayed wound healing rate remains a critical issue, especially in patients bearing large, open lesions [6].

Histopathological analyses of BU patient biopsies reveal unique features of *M. ulcerans* infections [7]. For instance, the coagulative necrosis seen in BU lesions displays a relative paucity of leukocytic infiltration close to the bacilli themselves [7, 8]. Instead, neutrophils are seen in the peripheral areas [9] and necrotic tissues often extend some distance from the site of mycobacterial colonisation [7, 10]. Both characteristics are caused by the lipid-like exotoxin produced by *M. ulcerans*, mycolactone [10, 11], which is synthesised by polyketide synthases encoded on a megaplasmid, pMUM001 [12].

Mechanistically, mycolactone causes cell cycle arrest in G_0_/G_1_ and subsequent progression to apoptosis after 48-72 hours, depending on cell type [10, 13]. Mycolactone can potentiate an integrated stress response via eIF2α phosphorylation and ATF4 activation [14, 15]. In addition, mycolactone has immunosuppressive effects on the maturation or functions of macrophages, dendritic cells and T cells [16–20]. All these activities can now be attributed to mycolactone’s inhibitory effect on Sec61 translocon [21, 22], which affects co-translational translocation of proteins into the endoplasmic reticulum (ER) [21, 23–26]. This blockade leads to protein translation in the wrong cellular compartment followed by their proteasomal degradation [21] and activation of autophagy pathways [14, 27]. Cells carrying mutations in the gene encoding Sec61α are resistant to mycolactone-driven cytotoxicity, supporting this as the primary target of mycolactone, at least at lower doses [14, 22, 23].

Mycolactone targeting of the Sec61 translocon is seen in various cell types, including endothelial cells, which regulate many physiological functions related to blood vessel homeostasis [21, 28]. Normally, endothelial cells form a monolayer that lines the lumen of blood vessels or form the capillaries. This semi-selective barrier controls the passage of small molecules and white blood cells into and out of the bloodstream [29, 30]. When blood vessels are injured, platelets rapidly attach to the damaged sites on the endothelium via its binding partner von Willebrand factor (vWF) [29, 31]. Secondarily to platelet plug formation, coagulation reinforces the plug by producing a fibrin mesh to hold it in place [29]. Fibrin clot formation can be triggered by both contact activation on a negative-charged, damaged surface or via the tissue factor (TF)-initiated extrinsic coagulation pathway. TF exposed from the sub-endothelium binds to blood-borne factor VIIa and other clotting factors, leading to thrombin activation and consequent processing of fibrinogen to fibrin [31]. These haemostatic pathways are regulated by the endothelium through its expression of specific molecules that function in anticoagulant pathways [30]. These include thrombomodulin (TM) and the endothelial cell protein C receptor (EPCR) that activate the protein C anticoagulant pathway [30, 32], tissue factor pathway inhibitor (TFPI) that supresses the factor VIIa-TF complex and free factor Xa activity [33], and heparan sulphate proteoglycans within the glycocalyx that bind antithrombin to directly inhibit thrombin [34].

We previously showed that mycolactone depletes thrombomodulin and the EPCR from primary human dermal microvascular endothelial cells (HDMECs) [28]. Reduced thrombomodulin staining was also seen in clinical specimens of BU where it was associated with fibrin deposition, itself strongly correlated with tissue necrosis [28]. While we did consider the dermis and subcutis separately, we acknowledged challenges in the analysis due to tissue heterogeneity. In order to overcome these challenges and to better understand the role and origin of fibrin deposition in BU lesions, originally reported in the mid-20^th^ Century, we performed immunohistochemical staining of serial tissue sections of BU patient skin biopsies for vascular and coagulation biomarkers. This more comprehensive analysis now shows an unexpected paucity of platelets, even in areas with extensive fibrin deposition, and widespread endothelial dysfunction. Fibrin deposition is explained by abnormal expression patterns of TF, implicating the involvement of extrinsic clotting pathway. The largest effect was driven by vessels that displayed TF staining at significant distance away from the basement membrane. In parallel experiments, our *in vitro* data showed that mycolactone increased the permeability of endothelial monolayers. Taken together our findings suggest that the vessels in BU lesions may become “leaky” during infection due to loss of the endothelial barrier integrity, and that this drives fibrin formation.

## Results

Immunostaining was performed on serial skin biopsy tissue sections from 8 untreated, laboratory confirmed BU patients (6 with ulcerative lesions and 2 with plaque, covering 3 WHO categories; Table 1 and S1 Fig). Repeat staining for several markers on different sections of the same punch biopsies in Ogbechi et al., 2015 [28] revealed that the pathology of Buruli ulcer is highly focal, even within these small 4mm samples of infected skin (S2A Fig). Hence, serial sections minimise drift throughout the biopsy sample and allow the tracking of individual vessels within the skin (S2B Fig) between the sections. The sections were stained in the following order: haematoxylin-eosin, fibrin, thrombomodulin (CD141), CD61, CD31 (Platelet endothelial cell adhesion molecule; PECAM-1), smooth muscle actin (SMA), Ziehl-Neelsen, TF and vWF. Regions of BU punch biopsy samples containing as many non-necrotic blood vessels as possible were defined by a pathologist prior to the start of analysis. This decision was made based on the number of blood vessels present that were positive for SMA or CD31 and whether there was any submacroscopic appearance of coagulative necrosis present in the corresponding H&E section. These “pre-defined” regions (see methods) are outlined in red in S1 Fig; nine areas were within the dermis and one area was located in the subcutis. Quantitative analysis concerned all identifiable vessels within these regions, regardless of whether nearby tissue displayed local signs of necrosis, in order to facilitate an unbiased analysis. Qualitative comparative analysis of the novel biomarkers not yet explored in BU (CD61, vWF and TF) was also performed across the whole biopsies, including areas that were highly necrotic. Acid fast bacilli were only visible in the sections from two of the patients (S2C Fig), therefore we do not report analysis of the coincidence of Ziehl-Neelsen staining further.

**Table 1.**
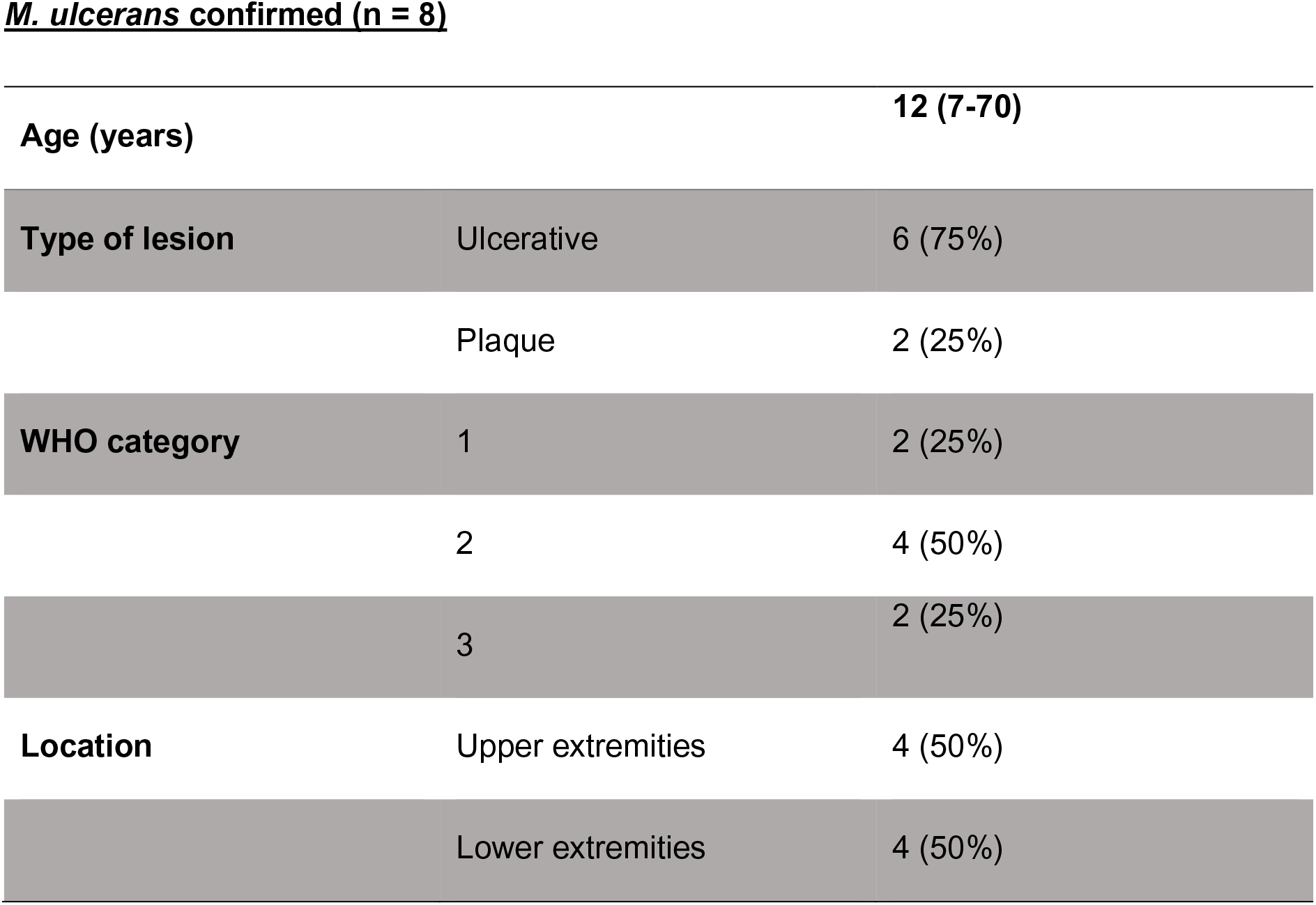
Clinical features of analysed Burui ulcer patients. Category 1: a single lesion < 5 cm in diameter; Category 2: a single lesion 5-15 cm in diameter; Category 3: a single lesion > 15 cm in diameter, multiple small lesions or facial lesions. Age is presented in median (with range) and the rest of data are n (%).

### Vasculopathy is evident in non-necrotic vessels of Buruli ulcer lesions

First, we closely analysed the phenotype of blood vessels in the defined regions of BU lesions using three well-known perivascular and/or endothelial cell markers: CD31, SMA and thrombomodulin. Since our previous studies showed that both thrombomodulin and, to a lesser extent, CD31 could be depleted by mycolactone exposure [28], we took a conservative approach and identified vessels which stained positively for SMA, or at least one of thrombomodulin and/or CD31. Using this approach, a total of 908 vessels were identified in the pre-defined regions of the eight patient samples (median of 92.5 vessels per patient sample, range 18-253).

When vessels were categorised according to the expression of these three markers (any level of positive staining), thrombomodulin presence was found to vary widely, regardless of the lesion type or WHO Category. All the vessels in the pre-defined region of one patient displayed were TM^+^ (Fig 1A), although for this patient the pre-defined region was very small and contained only 18 vessels (S1 Fig). In contrast, two patients had few vessels expressing thrombomodulin in the analysed areas, while other patients’ samples showed intermediate degrees of positive staining (Fig 1A). Overall, in the pre-defined regions containing non-necrotic vessels, only 299 out of 908 (32.9%) had normal staining (TM^+^ CD31^+^ SMA^+^). The remaining vessels lacked one or more of the expected vessel markers, with the most common phenotypes being TM^−^ CD31^+^ SMA^+^ (37.4%) and TM^−^ CD31^−^ SMA^+^ (18.7%). Some of the singly positive SMA structures could potentially be sweat glands with vessel-like morphology. However, even if this is the case for a proportion of these solely SMA^+^ structures, the data suggests a general vasculopathy in these BU lesions that precedes the emergence of coagulative necrosis.

**Fig 1.**
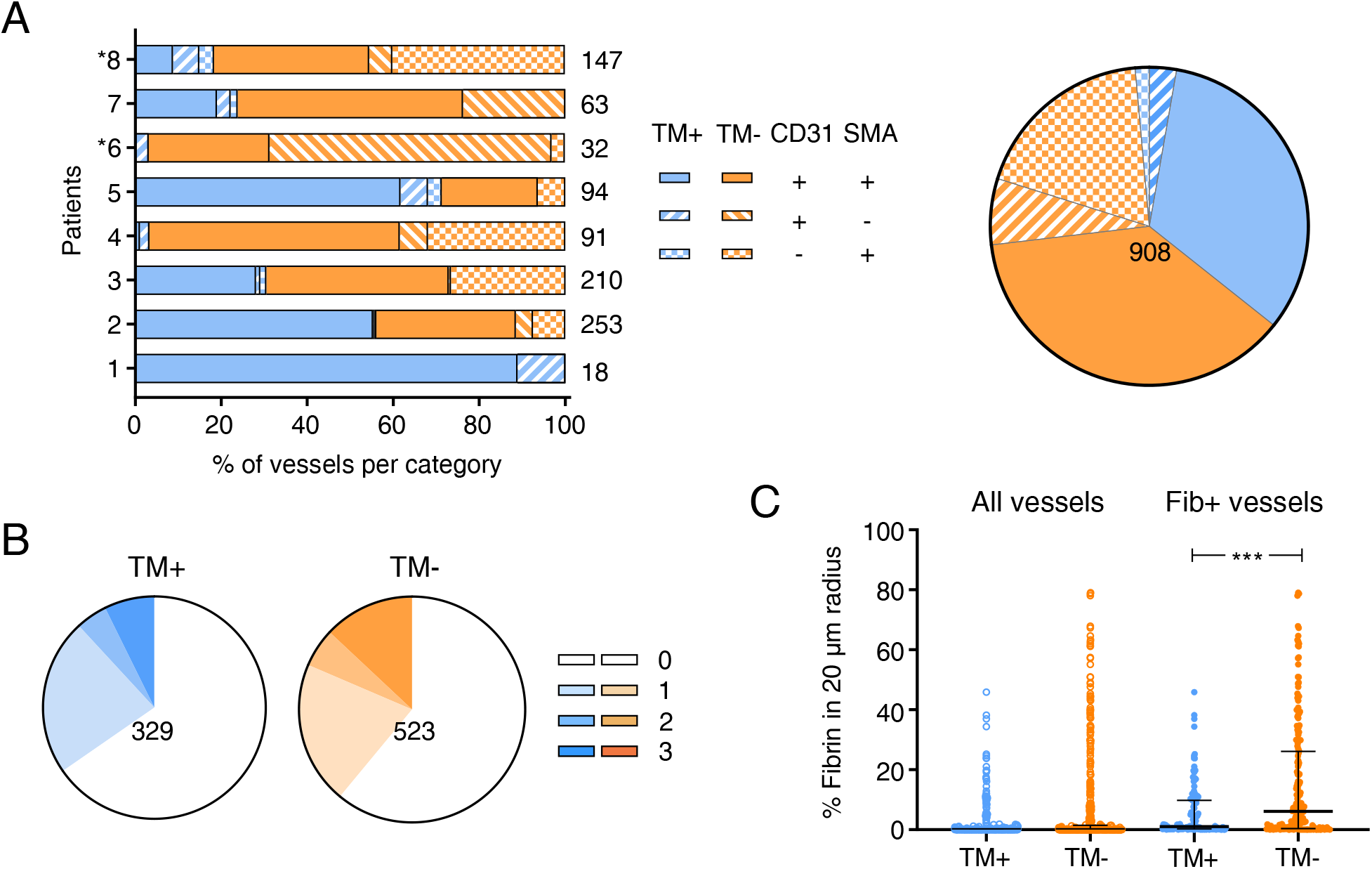
Thrombomodulin expression and fibrin deposition in and around analysed vessels. **A.** Vessels in pre-defined areas of punch biopsy samples lacking submacroscopic appearance of coagulative necrosis (see methods) of 8 BU patients (6 with ulcerated lesions and 2 with plaque lesions) were tracked by the endothelial cell marker CD31 and perivascular cell marker SMA, analysed and categorised according to the expression of TM, CD31 and SMA. The number of vessels in each category per patient was expressed as a percentage of the total number of the vessels counted for that representative area (listed right). Patients displaying plaque lesions are indicated with asterisks. The pie chart to the right shows the overall distribution of each category. **B and C.** The degree of fibrin deposition surrounding tracked vessels in A. **B** represents the distribution of fibrin scores of thrombomodulin-positive (TM^+^) and thrombomodulin-negative (TM^−^) vessels. Increasing fibrin scores corresponding to increasing extension of fibrin staining and are represented by stronger colour (0; no fibrin staining with 20 μm, 1-3 are fibrin staining in a <20, 20-30 and >30 μm radius, respectively). The total number of the vessels analysed is shown. **C.** The intensity of fibrin staining within a 20 μm area around each analysed vessel (TM^−^ or TM^−^) was quantified and expressed as percentage of the total analysed area for all and fibrin positive (Fib^+^) vessels. Data for individual vessels as well as median and interquartile range is shown. ***; *P* < 0.001.

In order to understand whether the previously described association between thrombomodulin depletion and fibrin deposition [28] is functionally linked, fibrin deposition was scored for each identified vessel in the defined regions, according to its degree of extension. Here, 852 vessels were trackable in analysed areas of tissue sections stained with fibrin, since not all vessels identified above could be found in subsequent sections. As reported previously [28], fibrin staining was seen across the biopsies, both in the defined regions and in highly necrotic areas but varying in intensity and extent for each patient (S1 Fig). The fibrin was mostly evident as tissue deposition surrounding, rather than in, blood vessels (298 vs. maximum 20 vessels in the defined areas).

Scoring of the fibrin disposition around 329 TM^+^ and 523 TM^−^ vessels (where 0 is no staining, and 3 is fibrin evident continuously for >30 μm from the vessel) showed thrombomodulin expression had no overall impact on fibrin levels (chi^2^ vs normal staining pattern *P* = 0.2007, Fig 1B). Hence, thrombomodulin depletion itself does not seem to be a good predictor of local fibrin deposition. However, while most vessels were fibrin-negative, in vessels lacking thrombomodulin expression the proportion with the highest fibrin score was increased compared to TM^+^ vessels (13.0 vs. 7.3%, Fig 1B). Moreover, the fibrin intensity within 20 µm of vessels lacking TM expression was much more variable than those where TM was detected (up to 79.0 vs. 45.8%, Fig 1C), suggesting the loss may aggravate fibrin formation in certain subgroup(s) of vessels. This is borne out by comparing the intensity of staining in fibrin-positive vessels, where TM^−^ vessels had significantly higher levels of fibrin within 20 µm.

### Primary haemostasis plays only a minor role in BU skin lesions, perhaps due to mycolactone-depletion of von Willebrand factor from endothelial cells

Since loss of thrombomodulin does not explain all the fibrin disposition seen in BU patient lesions, other pathways that could lead to fibrin formation were explored. First, we examined the localisation of molecules involved in primary haemostasis [30]. vWF is the primary component of Weibel−Palade bodies, secretory granules that are released from endothelial cells in response to activation or injury [35]. In healthy skin, vWF was present in endothelial cells lining blood vessels as expected [36] (Fig 2A1-2 and S3A Fig). As others have observed [37, 38], there is a relatively high stromal background for vWF staining due to its presence in serum. By contrast, in BU lesions, detection of endothelial vWF was frequently reduced (Fig 2B1, black arrowheads) or staining was seen in the intravascular space instead (Fig 2B1-2, black stars); rarely, vWF remained detectable in the endothelium (Fig 2B3, black arrow).

**Fig 2.**
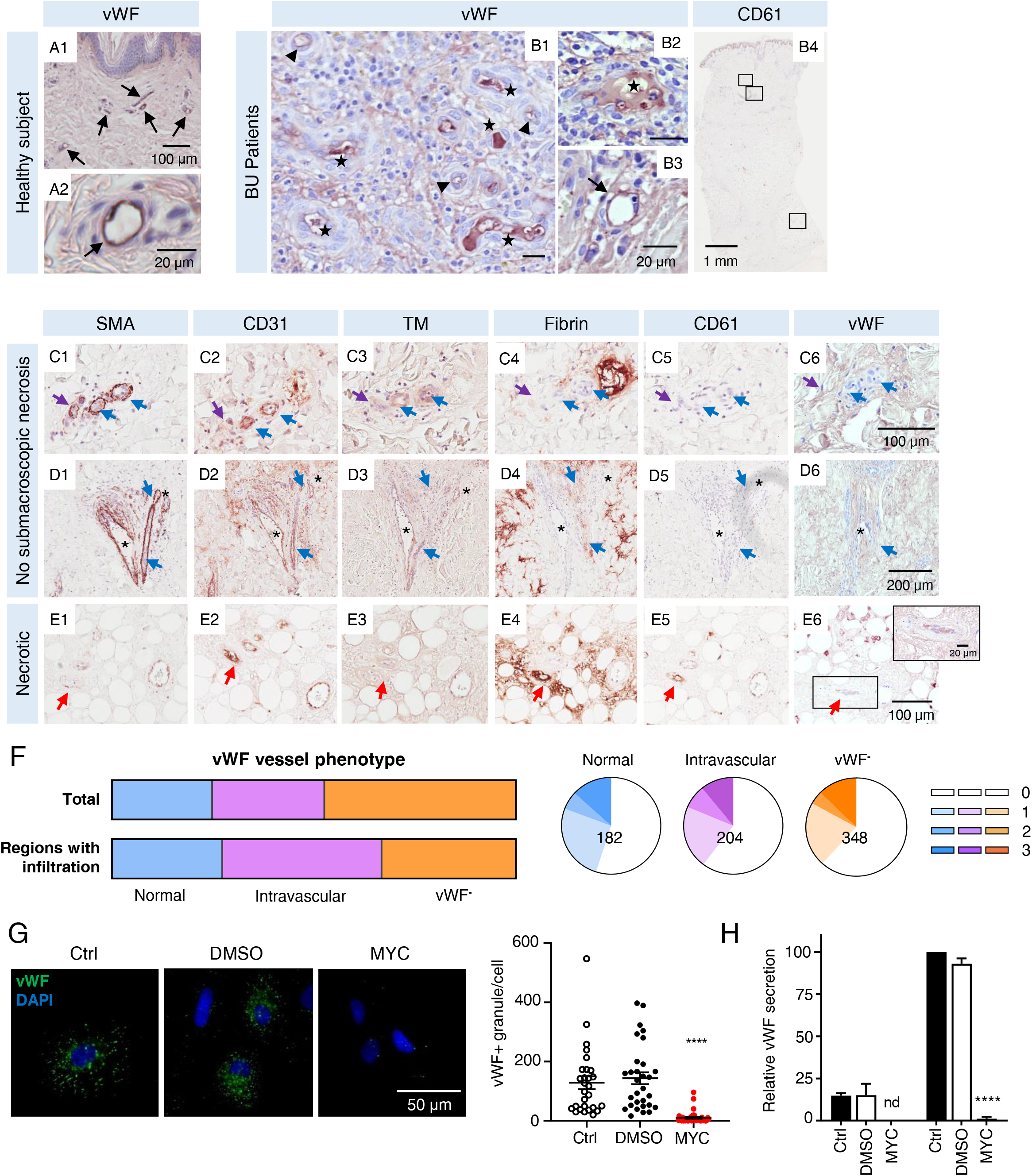
Endothelial von Willebrand factor is downregulated in response to mycolactone and redistributes in BU lesions. **A-E.** Histological sections stained with antibodies against vWF (A1-2, B1-3, C6, D6 and E6), platelet glycoprotein CD61 (B4, C5, D5 and E5), SMA (C1, D1 and E1), CD31 (C2, D2 and E2), TM (C3, D3 and E3) and fibrin (C4, D4 and E4) (positive staining in brown colour) and counterstained with haematoxylin in a healthy subject (A1-2) and BU patients (B, C, D and E). Scale bars as indicated. **A-B.** Examples of vessels with different phenotypes for vWF expression are indicated as follows; black arrows for normal expression, black arrowheads for reduced expression and black stars for vessels displaying intravascular vWF staining. Outlined regions in B4 showing distinct CD61 expression patterns in defined (the upper two; crop panels see C5 & D5) and in necrotic region (the bottom one; crop panel see E5). **C-D.** Examples of vessels with different phenotypes in pre-defined areas of BU punch biopsy samples lacking submacroscopic appearance of coagulative necrosis (see methods) are indicated as follows; purple arrows are SMA^+^CD31^−^TM^−^, blue arrows are SMA^+^CD31^+^TM^−^, and asterisks are SMA^+^CD31^+^TM^+^ vessels. **E.** An example of a CD61^+^ vessel displaying fibrin deposition in the necrotic subcutis is indicated with a red arrow. **F.** The overall distribution of vWF staining patterns in all patients (top panel) and vessels in immune cell-infiltrated regions (lower panel). The distribution of fibrin scores per vWF staining pattern (i.e. normal, intravascular and vWF^−^) is represented by pie charts with the total analysed vessel number shown. Increasing fibrin scores corresponding to increasing extension of fibrin staining are represented by darker colour (0; no fibrin staining with 20 μm, 1-3 are fibrin staining in a <20, 20-30 and >30 μm radius, respectively). **G & H.** Primary endothelial cells were treated with 10 ng/mL of mycolactone (MYC), 0.02% DMSO or untreated (Ctrl) for 24 hours. **G.** HUVECs were fixed, permeabilised and immunostained with anti-vWF antibody. vWF-containing granules are shown in green and nuclei stained with DAPI (blue). Scale bar = 50 µm. Scatter plot showing vWF-positive granules per cell in each condition. ****; *P* < 0.0001. **H.** HDMECs were treated with 2 U/mL thrombin to induce exocytosis of Weibel-Palade bodies. The concentration of vWF in supernatants was quantified by ELISA. nd, not detected.

One of vWF’s primary roles is to capture activated platelets, thus providing a rich negatively charged surface for amplification of the blood coagulation cascade leading to fibrin formation. Activated platelets express platelet glycoprotein IIb/IIIa (CD41/CD61), as their main receptor for vWF and fibrinogen, and here we examined the expression of CD61, also known as integrin β3 [39]. Surprisingly, considering the wide abundance of fibrin in the lesion, very little CD61 staining was seen in BU lesions (Fig 2B4 and S1 Fig), even in areas with coagulative necrosis. This suggests a minor role, if any for primary haemostasis in lesion formation.

To consider this more closely, we analysed the colocalisation of CD61 and vWF in BU patient vessels. In the defined regions (Fig 2C1-6, 2D1-6), CD61 and vWF were barely detectable in vessels, despite strong fibrin staining in the vicinity (Fig 2C4 and 2D4, blue and purple arrows). In vessels that were positive for of least one of SMA, CD31 or thrombomodulin, none stained positively for CD61. In necrotic regions in the subcutis, CD61 staining was occasionally seen in areas where fibrin deposition was evident, in vessels that were CD31^+^ and TM^−^, colocalising with vWF (Fig 2E1-6, red arrows).

We have already shown that platelet activation is not directly affected by mycolactone [28], and the CD61 presence did not correlate with vWF in non-necrotic vessels. Therefore, we postulated that this might be due to mycolactone-dependent depletion of vWF from endothelial cells, and there is some evidence to support this from these BU patient samples. Of 734 trackable vessels in defined regions in BU lesions, only 24.8% retained a normal expression pattern, with their endothelial monolayer stained positively for vWF (Fig 2B3 and 2F upper panel). Instead, nearly half (47.4%) had completely lost vWF staining (vWF^−^) (Fig 2B1, 2C6, 2D6 and 2F upper panel). Intriguingly, 204 vessels (27.8%) displayed vWF staining within the lumen of microvessels, in the intravascular space (Fig 2B1-2 and 2F upper panel). This pattern of staining was particularly evident in BU patient punch biopsies in which the pre-defined areas were heavily infiltrated with immune cells (S1 Fig, outlined in blue), representing 52.5% of such vessels (Fig 2F, lower panel). However, fibrin scores were similar across all three phenotypes of vWF expression in vessels (chi^2^ vs normal staining pattern *P* = 0.2882 and 0.1280 for intravascular and vWF^−^, respectively Fig 2F), suggesting that none of these phenotypes are directly related to fibrin formation.

Since vWF enters Weibel-Palade bodies via the secretory pathway and should be a Sec61-dependent protein [35], we reasoned that the widespread depletion of vWF in BU lesions might be related to the ability of the diffusible exotoxin mycolactone to inhibit the Sec61 translocon. Hence, we visualised vWF in cultured primary HDMECs and human umbilical vein endothelial cells (HUVECs) by immunofluorescence (Figs 2G and S3B Fig). Although the amount of vWF-positive granules (presumptive Weibel-Palade bodies) varied from cell to cell, these were greatly reduced after 24 hours exposure to mycolactone (Fig 2G). Furthermore, when we quantified the level of vWF in conditioned medium from HDMECs by enzyme-linked immunosorbent assay (ELISA), both basal and thrombin-induced vWF secretion was almost completely abolished following mycolactone exposure (Fig 2H).

### Extravascular tissue factor within BU lesions correlates to pathogenic fibrin deposition

Another potential trigger for fibrin generation is activation of the extrinsic coagulation cascade, initiated by TF. In the skin of healthy individuals, normal TF staining is found only in the epidermis and the adventitia of larger (but not smaller) blood vessels as expected (Fig 3A1-2 and S4A Fig). This highly restricted expression pattern is vital to maintain intravascular fluidity, due to its ability to rapidly catalyse activation of the coagulation cascade by binding Factor VII/VIIa. In BU patient lesions, TF staining was not confined to the sub-endothelium and was instead seen within the tissues in a complex, highly variable pattern between patients. For example, in one patient, TF was present at the edge of damaged area (S4C Fig) where extravasated erythrocytes had accumulated. Abnormal TF staining fell into one of four main categories. First, staining outside of the vessel basement membrane (BM) of larger vessels (Fig 3B5 and 3C5, red arrows). Second, in presumptive infiltrating cells, either demonstrating granular intracellular staining (Fig 3D5 and S4B Fig, black arrows) or intensively throughout each cell (Fig 3E5, black arrows). This pattern of staining was seen in both necrotic and in defined regions, and in the latter case was in regions that appeared to contain eosinophils based on Haematoxylin and eosin (H&E) staining (Fig 3E3, black arrowhead). Third, TF staining was found within the endothelial cells lining smaller vessels (Fig 3F5, orange arrow), which do not normally express this receptor. Lastly, some vessels contained some structures within the vessel lumen that could not be identified, yet stained strongly for TF (Fig 3G5, purple arrows). Approximately one quarter of vessels with this phenotype were in infiltrated regions (Fig 3G3).

**Fig 3.**
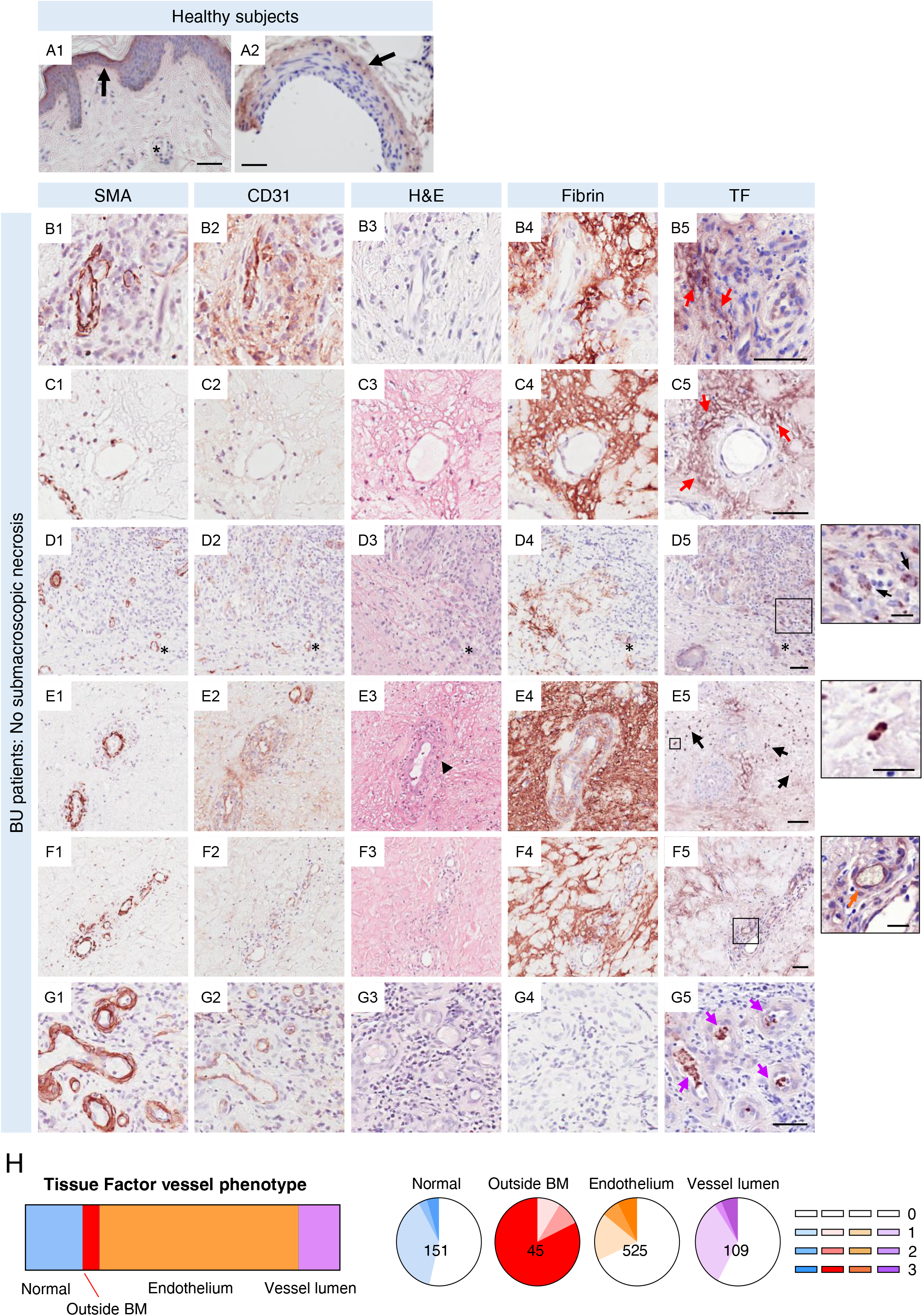
Tissue factor expression is altered in BU patient skin lesion. **A-G.** Histological sections were stained with anti-tissue factor (TF) antibody (A1-2, B5, C5, D5, E5, F5 and G5), anti-SMA antibody (B1, C1, D1, E1, F1 and G1), anti-CD31 antibody (B2, C2, D2, E2, F2 and G2), eosin (B3, C3, D3, E3, F3 and G3) or anti-fibrin antibody (B4, C4, D4, E4, F4 and G4) (positive staining in a brown colour) and counterstained with haematoxylin in healthy subjects (A) or in pre-defined areas of BU punch biopsy samples lacking submacroscopic appearance of coagulative necrosis (see methods) (B-G). Scale bar = 50 µm (20 µm in the crop panels of D5, E5 and F5). **B & C.** Red arrows indicate regions of TF staining distant from the vessel basement membrane (BM). **D.** Asterisks label the same vessel, black arrows in the crop panel of D5 indicate examples of cells containing punctate structures staining strongly for TF. **E.** Black arrows indicate examples of cells staining intensively for TF throughout the cells, in regions where cells stained intensively for eosin in the H&E stain (black arrowhead). **F.** An orange arrow in the crop panel of F5 indicates an example of the endothelium of a small vessel stained positively for TF. **G.** Purple arrows indicate examples in a region where multiple small vessels contained unidentified structures stained positively for TF. **H.** The overall distribution of vessel phenotypes for TF staining across 8 BU patients. The distribution of fibrin scores associated with each phenotype are represented as pie charts. Increasing fibrin scores corresponding to increasing extension of fibrin staining are represented by stronger colour (0; no fibrin staining with 20 μm, 1-3 are fibrin staining in a <20, 20-30 and >30 μm radius, respectively). The total number of the vessels analysed is shown.

With respect to TF-positive presumptive-infiltrating cells, these were observed within tissue rather than close to vessels. Granular intracellular staining was seen in 5 of the 8 BU patient biopsies, almost exclusively in the defined regions where they were between 2.2-89.9 μm away from the vessel BM. In most of these patients (4 out of 5) it occurred in areas displaying fibrin deposition (compare Figs 3D4 and 3D5). Infiltrating cells displaying intensive TF-positive staining throughout each cell body were seen in 6 of the 8 BU biopsies and always in regions of fibrin accumulation. This phenotype was more common in the necrotic subcutis (5 patients). In the one patient where it was seen in a defined region, the cells were 17.1-120.9 μm away from the vessel BM.

Vessels in defined regions were tracked and classified according to their TF expression patterns. Of 830 trackable vessels, 18.2% showed normal TF expression in the adventitia, 45 (5.4%) showed TF staining outside of the vessel BM, within the skin tissue, 63.3% displayed abnormal TF staining in the vessel endothelium, and 13.1% showed TF within the vessel lumen (Fig 3H). All abnormal expression patterns were associated with an increased chance of having the highest fibrin score. While this was not significantly different for vessels where TF staining was seen in the lumen (chi^2^ vs normal staining pattern *P* = 0.5901), it was distinct in vessels where the endothelium was TF^+^ (chi^2^ vs normal staining pattern *P* = 0.0014). Most strikingly, whilst being relatively rare in the defined regions, vessels that displayed TF staining outside of BM were significantly more likely to have fibrin deposited in the surrounding tissue, (chi^2^ vs normal staining pattern *P* < 0.0001, Fig 3H) and more likely to have fibrin that extended further from the vessel (82.22% exhibiting the highest fibrin score). Notably the coincidence of fibrin and TF outside the BM was often found where there were local signs of tissue necrosis by H&E staining (e.g. Fig 3C3), suggesting that fibrin-deposition may cause result directly in necrosis, presumably due to ischaemia.

### Mycolactone reduces the expression of tissue factor pathway inhibitor in endothelial cells

The interaction between TF and the 50 kDa blood-borne factor VIIa, and the subsequent triggering of the coagulation cascade leading to fibrin formation is normally prevented by a functional endothelial barrier which segregates bloodstream from the TF-containing subendothelium [30]. Normally, TF/factor VIIa activity is suppressed by TFPI, which is mostly expressed by vascular endothelium and megakaryocytes [33]. Endothelial cells predominantly express the membrane-associated TFPI splice variant lacking both the C-terminus and the third Kunitz-type inhibitory domain (isoform β), but also express full-length TFPI (isoform α). Upon stimulation or heparin infusion, TFPIα, bound to cell-surface glycosaminoglycans through its C-terminus, can be secreted rapidly [40]. As fibrin deposition was found in BU affected tissues and correlated to TF seen outside of vessel BM, we wondered whether TFPI expression might be affected by mycolactone. As we were unable to test this in the biopsies themselves, we investigated this *in vitro*. TFPI was rapidly depleted from HDMECs in response to mycolactone exposure with a decrease detectable after 8 hours by Western blot (Fig 4A); given that TFPIα and β have different levels of glycosylation and migrate with similar molecular mass on sodium dodecyl sulfate polyacrylamide gel electrophoresis [41], we couldn’t distinguish between these isoforms. Quantification of this data demonstrated a 57.4% reduction of total TFPI after 8 hours of mycolactone exposure and further to 88.3% after 24hours (Fig 4A). A similar decline was seen in conditioned medium, where the amount of TFPI secreted by HDMEC over 24 hours reduced from 4.59±0.18 to 0.74±0.13 ng/mL (Fig 4B).

**Fig 4.**
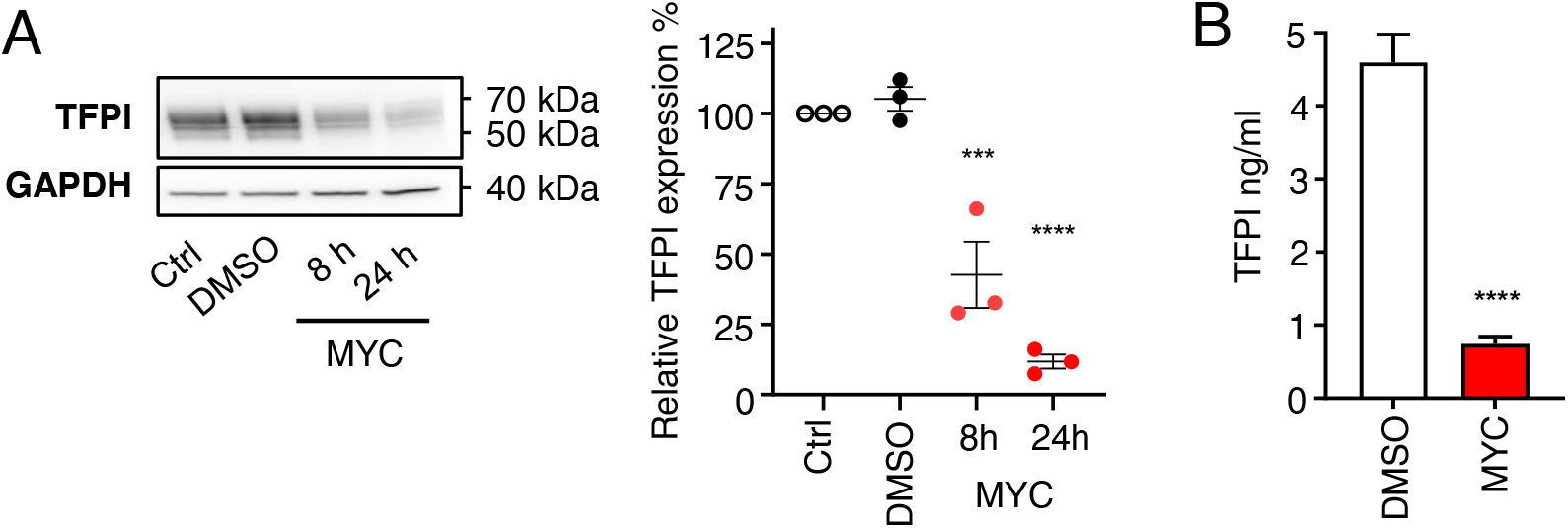
Mycolactone reduces the expression of tissue factor pathway inhibitor in endothelial cells. **A.** HDMECs exposed to 10 ng/mL of mycolactone (MYC) for the indicated times, 0.02% DMSO or untreated (Ctrl) were lysed and subjected to immunoblotting. TFPI immunoblot intensity was normalised according to GAPDH and untreated controls. Data from 3 independent experiments are presented (mean ± SEM). **B.** Supernatant was collected from cells treated as above for 24 hours, cell debris removed and TFPI was quantified by ELISA. Values represent the mean of three independent experiments ± SEM. ***, *P* < 0.001; ****, *P* < 0.0001.

### Mycolactone-dependent vascular permeability as a potential driver of coagulopathy in Buruli ulcer lesions

Taken together the data from BU punch biopsies suggests that the expression of haemostatic markers on the vessel endothelium is not a reliable predictor of fibrin deposition close to that vessel. Instead, fibrin deposition in the tissue seems to arise locally and driven by TF expression that is uncontained within the stroma. To examine how uncontained TF may impact fibrin deposition in such a distinct phenotype, the staining intensity of TF and fibrin were quantified and the distance of the TF signal from the vessel BM were measured in these same vessels. There was no correlation between fibrin and TF staining intensity within 20 µm (*r* = - 0.1623, *P* = 0.2869) (Fig 5A). However, there was a positive correlation (*r* = 0.2981, *P* = 0.0467) between fibrin intensity and the distance of TF^+^ signal from the same analysed vessel BM (Fig 5B).

**Fig 5.**
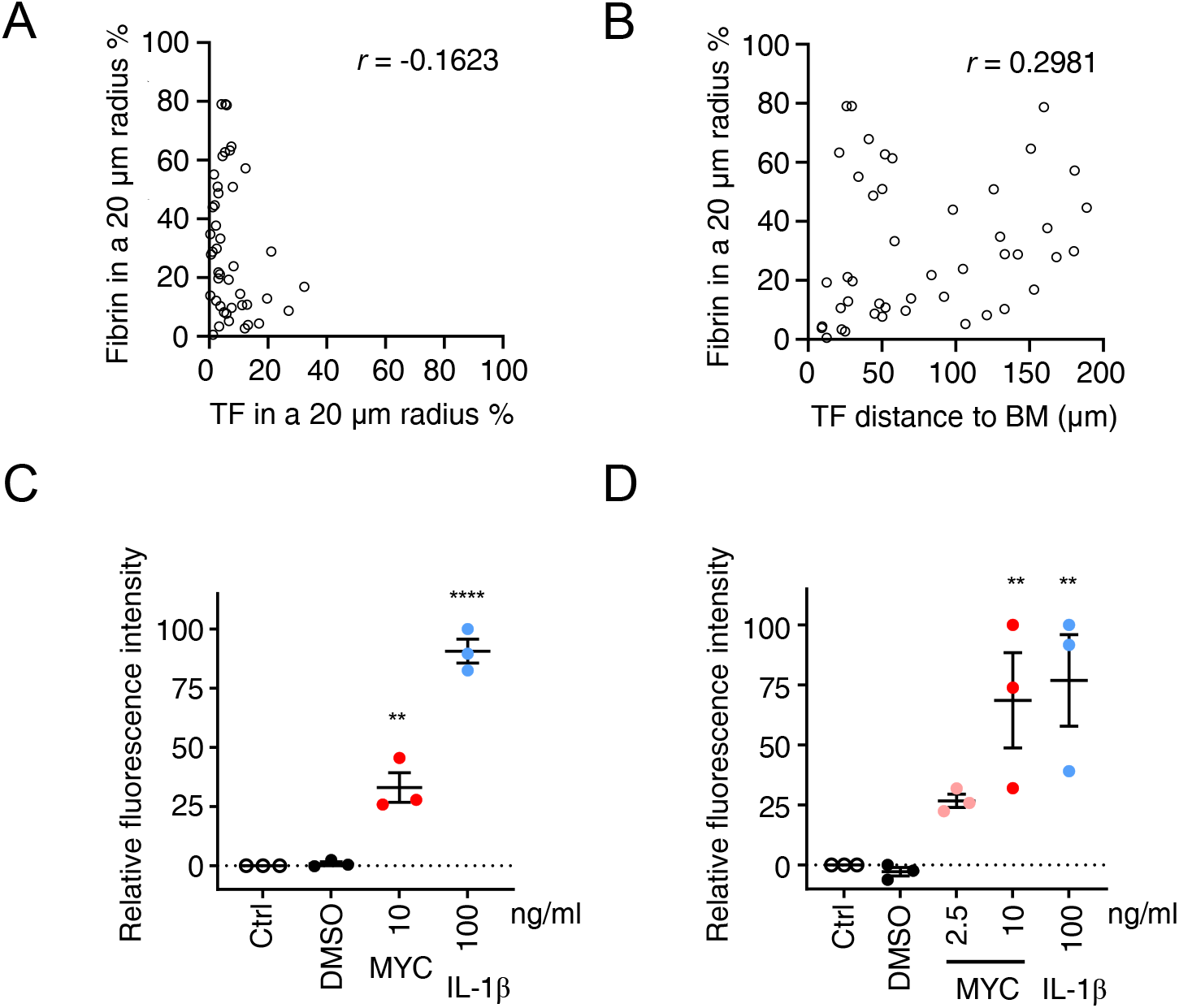
Extravascular tissue factor (TF) is the primary driver of fibrin deposition in BU lesions, potentially driven by an increase in vascular permeability. **A & B.** Correlation between the abundance of fibrin within a 20 μm radius of each vessel and the abundance of tissue factor (TF) in the same radius (**A**) or the distance the TF^+^ signal extended from the vessel basement membrane (BM) (**B**) for 45 trackable vessels. *r* is Spearman’s rank correlation coefficient for each data set. **C & D.** Permeability of HDMEC (**C**) or HDLEC (**D**) monolayers, on inserts with 0.4 and 1 μm pores respectively, that remained untreated (Ctrl), or were exposed to 0.02% DMSO, 2.5 or 10 ng/mL mycolactone (MYC), or 100 ng/mL IL-1β for 24 hrs. Fluorescence intensity of FITC-dextran in the receiver wells was measured and presented as a % where 100% is the value obtained from transwell lacking a cell monolayer, and 0% is untreated control wells. Values represent the mean of three independent experiments ± SEM. **, *P* < 0.01; ****, *P* < 0.0001.

Generation of fibrin within tissue, a substantial distance away from vessels, resulting from extrinsic coagulation pathway activation would require large plasma glycoproteins to gain access to the stromal compartment. We therefore asked whether mycolactone might induce changes to vascular permeability that could lead to a breakdown of vessel integrity *in vitro*. Hence, we evaluated the effect of mycolactone on the permeability of HDMEC monolayers to 70 kDa FITC-labelled dextran in a transwell system. In line with our hypothesis, exposure to 10 ng/mL mycolactone for 24 hours increased the permeability of the monolayer to 33.1±6.2% of the value seen when cells were completely absent from the transwell (Fig 5C), equivalent to around one-third of that seen with interleukin-1β (IL-1β), a known inducer of vascular permeability [42].

We also wondered whether this mycolactone-dependent increase in permeability might extend to the lymphatic system, since BU can sometimes have an oedematous presentation. In human dermal lymphatic endothelial cells (HDLECs), a subpopulation of endothelial cells that actively participate in fluid balance and transport [43, 44], this phenotype was even more apparent. HDLECs and HDMECs had comparable half maximal inhibitory concentration to mycolactone (around 0.67 ng/mL, S5 Fig), but the response of lymphatic endothelial cells to mycolactone was more pronounced. Exposure to 10 ng/mL mycolactone for 24 hours increased the permeability of the monolayer to 68.6±19.8% of the value seen in empty wells (Fig 5D). In the same time window, 2.5 ng/mL mycolactone could potentiate this effect but to a lesser degree (26.7±2.8%, Fig 5D).

### IL-1β aggravates mycolactone-driven endothelial dysfunction

Recent work showed that IL-1β can be induced by mycolactone-containing microvesicles and is found in *M. ulcerans*-infected tissues [45]. Since IL-1β has been long-known to repress thrombomodulin expression in endothelial cells by a transcriptional mechanism [46], and also induces vascular permeability [42], we wondered whether the presence of this alarm cytokine might exacerbate the effect of mycolactone on the microvasculature. First, we addressed whether mycolactone’s ability to supress thrombomodulin expression in primary endothelial cells might be influenced by IL-1β, by exposing cells to a range of concentrations of mycolactone and/or IL-1β. As expected, exposure to either 10 ng/mL IL-1β or 2.5 ng/mL mycolactone alone for 24 hours led to a significant depletion in thrombomodulin protein level in both HDMECs (Fig 6A) and HDLECs (Fig 6B). Similar to our observations regarding vascular permeability, lymphatic endothelial cells were more sensitive to both stimuli, with >50% reduction in the presence of 0.6 ng/mL IL-1β (*P* = 0.0132) or 1.25 ng/mL mycolactone (*P* = 0.0004) (Fig 6B). Remarkably, this downward trend was more evident when the endothelial cells were co-exposed to non-saturating amounts of both agents (Fig 6A and B). For example, in HDMEC, while each stimulus alone resulted in ∼55-75% depletion of thrombomodulin expression, it was barely detectable in endothelial cells exposed to both 2.5 ng/mL mycolactone and 10 ng/mL IL-1β. In HDLEC, the combination of 0.625 ng/mL mycolactone and 0.6 ng/mL IL-1β suppressed thrombomodulin expression to the same extent as 10 ng/mL IL-1β. These effects appear to be additive rather than synergistic.

**Fig 6.**
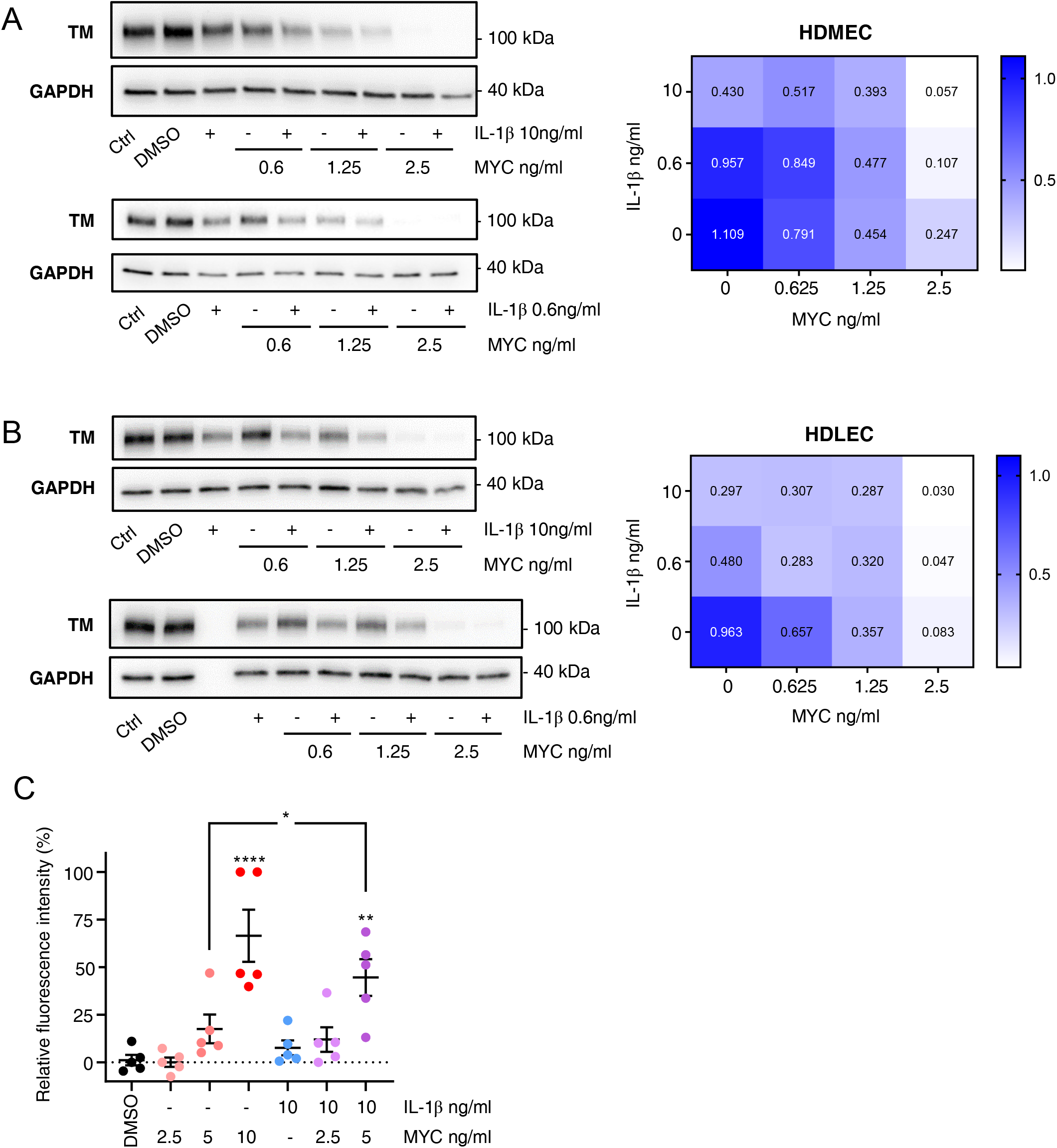
IL-1β aggravates mycolactone-driven endothelial dysfunction. HDMECs (**A**) or HDLECs (**B**) were exposed to different concentrations of mycolactone (MYC), and/or IL-1β for 24 hours, then lysed and subjected to immunoblotting. TM immunoblot intensity was normalised according to GAPDH and expressed relative to untreated controls. Mean expression level from 3 independent experiments are presented as a heatmap where increased TM loss is represented by a paler colour. **C**. Permeability of HDMEC monolayers on inserts with 1 µm pores was quantified after exposure to mycolactone (MYC) and/or IL-1β for 24 hours. Fluorescence intensity of FITC-dextran in the receiver wells was measured and presented as a % where 100% is the value obtained from transwell lacking a cell monolayer, and 0% is untreated control wells. Values represent the mean of five independent experiments ± SEM. *, *P* < 0.05; **, *P* < 0.01; ****, *P* < 0.0001.

Since the additive effect was particularly evident in HDMECs, we investigated whether it might also influence vascular permeability in these cells. Here, we used a lower dose of IL-1β, which only slightly increased the permeability of monolayer after 24 hours (7.66±3.89% of the value seen in an empty well, Fig 6C). Mycolactone had a dose-dependent effect on vascular permeability as before, and the additive effect of IL-1β was evident when there was 5 ng/mL mycolactone, increasing the permeability significantly from 17.58±7.57% to 44.62±9.65% (*P* = 0.0021 vs. DMSO; *P* = 0.0268 vs. mycolactone alone). This data suggests that even low amounts of IL-1β in infected tissues might have a further negative impact on haemostatic status and endothelial monolayer integrity where trace amounts of mycolactone are present.

## Discussion

Coagulation is often considered as a frontline host defence response to infection due to trapping of invading bacteria clots [47]; however, some pathogens develop strategies to bypass or utilise clot architecture to gain access to host resources. For example, *Streptococci* generate streptokinase, a plasminogen activator mimic, that promotes fibrinolysis and thus has a positive impact on bacterial metastasis [48]. Coagulase-positive *staphylococci*, on the other hand, produce coagulase, which rapidly depletes fibrinogen from the plasma and blunts the phagocytic activity of neutrophils [49]. Malaria-infected erythrocytes cause the loss of EPCR and thrombomodulin in brain vessels, leading to subsequent fibrin deposition within the microvasculature [50], which impacts the severity of cerebral malaria [51]. In the case of BU, the skin lesions exhibit large-scale coagulative necrosis in the subcutaneous regions, a distinct pathological phenotype, which has been long associated with the biological effects of the exotoxin mycolactone as it diffuses through the infected tissue [52].

Cumulative reports with *in vitro* or *in vivo* assays have addressed how cell death can be potentiated in response to mycolactone, either directly via the activation of caspase-3, caspase-9 [53] or Bcl-2-like protein 11 [54] with resultant apoptosis, and/or via an integrated stress response leading to ATF4 activation [14, 15]. These pathways may also involve disruption of actin cytoskeleton [55]. More recently an indirect mechanism was proposed, via mycolactone’s Sec61-dependent influence on coagulation control and resultant tissue ischemia [28]. However, to what extent coagulation is dysregulated in BU has been unclear. This present work demonstrates several abnormal phenotypes for haemostatic proteins in the dermal microvasculature of BU patients that includes: 1) loss of key endothelial proteins such as the endothelial adhesion molecule CD31, the anticoagulant thrombomodulin, and the platelet binding partner vWF, and 2) highly abnormal expression patterns of TF that suggest a loss of containment. These changes likely underpin the pathogenic fibrin deposition seen in the skin lesions and imply that infection with *M. ulcerans* may cause a localised, disturbed haemostatic status. This conclusion is reinforced by *in vitro* data using primary endothelial cells, in which we show that mycolactone can potentiate vascular and lymphatic hyper-permeability. Moreover, mycolactone limits the expression, and secretion of, vWF and TFPI.

Mechanistically, a combination of mycolactone’s action at the Sec61 translocon, and increased local concentration of IL-1β, likely explains the endothelial dysfunction seen in BU. Like the single-pass type I membrane proteins thrombomodulin and CD31 [28], the secretory proteins vWF and TFPI require Sec61-dependent co-translational translocation into the ER lumen to complete their biosynthesis, processes that are inhibited by mycolactone [24, 25]. In *in vitro* model systems, the mycolactone concentration is uniform, whereas in BU skin lesions the situation is more complicated, but where mycolactone is present at high enough concentrations, depletion of these Sec61-dependent molecules should also occur as the process is conserved amongst diverse cell types. The variations in endothelial marker expression seen in different specimens and sections from the same specimen may therefore reflect local variations in mycolactone concentration. However, while there is much evidence for mycolactone within BU lesions [10, 56], these currently lack the spatial resolution to understand exactly how much mycolactone is present at specific sites.

It is clear from our data that in the complex structure of the disordered skin in BU that there was less than expected correlation between loss of endothelial transmembrane proteins, based on Sec61 inhibition alone. For instance, while many vessels lost both thrombomodulin and vWF, some had lost expression of one marker but not the other. Since it was recently shown that IL-1β is present in BU skin lesions also with variable intensity across biopsy samples [45], we tested the hypothesis that this may add an additional layer of complexity in the *in vivo* setting. We showed that mycolactone and IL-1β have an additive effect *in vitro*, suggesting that the variation in marker expression could be caused by the cumulative effect of local mycolactone and IL-1β. In this context, it is also worth noting that the distance between our serial tissue sections for these markers is 24 μm inclusive and given the focal nature of the staining for the same markers across the section (see Fig S2A), this could also be a contributing factor.

These BU patients showed widespread changes to the localisation of vWF in skin punch biopsies, with only a minority of vessels in defined regions displaying a normal expression pattern. In healthy tissue, vWF is expressed solely in endothelial cells, and is found contained within organelles called Weibel-Palade bodies (WBPs), which also contain a variety of other components regulating inflammation as well as vascular modulators [57]. Endothelial cells respond rapidly upon activation by a variety of so-called secretagogues to exocytose their WBP, upon which the vWF forms multimeric strings of molecules whose normal function is to capture platelets [58]. It is possible that WBP exocytosis explains the pattern we observed, particularly in immune cell infiltrated regions, where vWF was lost from endothelial cells and instead observed in the adjacent intravascular space. This pattern of expression has been previously reported in the dermal vasculature of patients with malignant melanoma [59] as well as alveolar septa and large blood vessels of patients with malaria-associated acute respiratory distress syndrome [37]. In melanoma vasculatures, CD42^+^ platelet aggregates were also seen bound to intravascular vWF fibres [59], due to an endothelial glycocalyx shedding-dependent process [60]. We did not observe platelet aggregates in these vessels in our BU patients, although we cannot completely rule out that this is a technical limitation of the antibody used to detect them [61].

Another haemostatic factor with a grossly abnormal expression pattern was the extrinsic pathway initiator TF, which plays a critical role in both fibrin generation and wound repair [62]. Exposure of TF to blood is sufficient to initiate clotting, due to its cofactor activity in the activation of factors IXa and Xa by factor VIIa [63]. Consequently, TF expression in the skin is normally restricted to the epidermis and the adventitia surrounding larger vessels as a so-called “haemostatic envelope” [63], and its activity is usually tightly regulated by an inhibitor, TFPI [33]. Changes in TF expression have been reported in many inflammatory thrombotic conditions [62, 64], as well as in infections such as tuberculosis [65, 66]. Myeloid cells do not normally express TF, but during *M. tuberculosis* infection, TF expression is observed in macrophages along with conspicuous fibrin deposition within granulomas [66, 67]. Mice genetically altered to have low TF expression [65] or TF deficiency in myeloid cells [67] do not show these changes.

Whether the endothelium can be a cellular source of the TF in pathogenic conditions remains highly controversial [62, 68]. It is well-established that inflammatory agents such as lipopolysaccharide, oxidized low density lipoprotein and IL-1β can induce TF mRNA and protein expression in monocytes and macrophages both *in vitro* and *in vivo* [69–71]. On the other hand, while endothelial cells can be induced to express TF *in vitro* in response to lipopolysaccharide and IL-1β [42, 72, 73] the evidence that this takes place *in vivo* as part of a pathophysiological process is not so clear. For instance, endothelial-specific TF-knockout mice display little influence over pathogenesis in a range of disease models [62, 74]. Much evidence supports a view that procoagulant TF always arises via activated mononuclear cells and that the detection of TF in other cells is a consequence of microparticle uptake [62, 75]. Indeed, early reports that eosinophils could express TF following activation [76] were more recently shown to be explained by their uptake of monocyte-derived microparticles [68]. While this is a highly complex and also controversial area, many cell types including endothelial cells, platelets, and monocytes/macrophages have all been shown to be capable of producing TF-bearing microparticles *in vitro*, but monocyte-derived microparticles have been shown to be the most thrombogenic [77]. As well as providing the extrinsic clotting pathway initiator TF, microparticles are also thought to promote coagulation by providing a rich negatively charged surface for the amplification of the coagulation cascade at their location [62]. The presence of circulating TF-bearing microparticles has been reported in patients with cancer-associated thrombosis [78], venous thromboembolism [79], cardiac bypass surgery [80], Behçet’s disease [81] and sepsis [82].

In the BU patients we investigated, epidermal expression of TF was not altered, but within the dermis and subcutis extremely disordered TF expression was observed. In stark contrast to healthy skin, even in the pre-defined regions containing non-necrotic vessels, TF was detected in the endothelial cells of >60% of tracked vessels, as well as in infiltrating cells in five patients with either granular, or one patient with whole-cell, staining. This finding, reminiscent of inflammatory thrombotic conditions, such as sepsis and atherosclerosis [62, 64, 68], was surprising because of mycolactone’s well-characterised immunosuppressive and anti-inflammatory effects [56]. Once again, IL-1β may play an important role here. We speculate that the TF^+^ endothelial cells and presumptive eosinophils we observed could have arisen by uptake of TF-bearing microparticles induced by IL-1β within the lesions. It is unlikely that TF in the endothelium has much influence over disease progression in these BU patients, as fibrin deposition was little altered around these vessels even when the anticoagulant thrombomodulin was also lost from the endothelial cell surface. On the other hand, we observed TF^+^ cells (presumably eosinophils) in necrotic and, more rarely, regions of BU lesions containing plentiful non-necrotic vessels, where fibrin was seen intensively in tissues. Notably, eosinophils have also been reported as the cellular source of TF seen in another skin condition, chronic urticaria [83]. It is also possible that some of the infiltrating TF^+^ cells we observed are activated macrophages that have been induced to express TF. *M. ulcerans* infection is known to include a phase where macrophages are transiently infected with bacteria [84], and the death of these cells due to the cytotoxic effects of mycolactone could conceivably be another source of microparticles. Whether microparticles of any origins play an important role in BU will be the subject of future investigations.

The strongest association we observed with fibrin deposition was diffuse TF staining extending away from larger vessels, up to 200 μm into tissue. A key question that arose was; how do TF and the other plasma clotting factors required to produce thrombin and consequently process fibrinogen to fibrin access these sites? Using an *in vitro* vascular permeability assay, we showed that mycolactone potentiates the passage of 70 kDa-size dextran through monolayers of both HDMECs and HDLECs. Given that adhesion molecules such as CD31 [85], thrombomodulin [86, 87] and vascular endothelial cadherin are essential regulators for endothelial tight junctions, and these proteins are downregulated by mycolactone [28], a disturbed endothelial physical barrier between plasma and tissue could be an unavoidable phenotype in BU. A leaky vasculature is well-established to result in increased immune cell invasion, as well as allowing plasma components and fluids to enter tissue [88, 89]. Hence, the disrupted physical barrier may initiate a clotting cascade as blood-borne clotting factors engage with their sub-endothelium located binding partners or activators [90].

Inflammatory cytokines are known to affect both microvascular and lymphatic endothelial barriers and augment vascular permeability [88, 91], and this can cause tissue oedema in many disease conditions [29, 88, 91]. In BU, oedematous forms are seen in around 5% of patients [92]. Further work will be required to establish how early in the infection process vascular permeability increases, and to understand whether it has a direct impact on *M. ulcerans* growth. Mycolactone’s effect on vascular permeability *in vitro* occurred much earlier (within 24 hours) than its cytotoxic effects (more than 72 hours in HDMECs) and at an extremely low dose (10 ng/mL), even lower in the presence of small amounts of IL-1β. Furthermore, while increased vascular permeability is a common strategy used by innate host defence systems to fight against invading microorganisms, many pathogens also target cell-cell junctions in order to cross the barrier and colonise in tissues. Interestingly, *Mycobacterium marinum*, a close genetic relative of *M. ulcerans*, targets vascular integrity to aid its multiplication in granulomas [89].

Taken together, both our cell-based studies and histopathological analysis in patient specimens illustrate a hypercoagulative microenvironment that develops during BU disease progression. It seems likely that the fibrin deposition seen in BU skin punch biopsies is not driven by platelet aggregation; instead, it is likely to be the consequence of factor VIIa engaging with sub-endothelial TF as the endothelial cell monolayer integrity is disturbed, exacerbated by the loss of its natural inhibitor TFPI. We have not yet analysed any potential contribution of the intrinsic clotting pathway; therefore, this cannot be excluded. A correct balance of the TF-driven coagulation cascade and subsequent fibrin formation is critical in wound healing [93]. Hence, application of anticoagulants along with antibiotics could help neutralise the pro-thrombotic phenotype seen in BU patient skin lesions, and ultimately improve healing rate. Of note, complementary anticoagulant heparin alongside standard anti-tubercular antibiotics treatment has been previously employed to treat a case of facial BU. In this patient, facial oedema was reversed by heparin intravenous injection [94], suggesting lowering haemostatic status may attenuate BU disease progression.

## Materials and Methods

### Ethics statement

Ethical approval for analysing BU patient punch biopsies was obtained from the Ethikkommission beider Basel, Basel, Switzerland and the provisional national ethical review board of the Ministry of Health Benin (N IRB00006860) as well as from the Cameroon National Ethics Committee and the Ethics Committee of the Heidelberg University Hospital, Germany (ISRCTN72102977). A favourable ethical opinion for analysing normal human skin was given by Faculty of Health and Medical Science Ethics Committee of the University of Surrey (1174-FHMS-16). The normal human skin samples were collected by the Whiteley Clinic, Guildford, Surrey or were purchased from AMS Biotechnology. Written informed consent was obtained from adult patients or the guardians of child patients. The research related to human tissues complies with the ethical processes of University of Surrey.

### Histological analysis of human skin samples

Buruli ulcer punch biopsies (4 mm) were collected previously [95] and reanalysed for the current study. The biopsies from 4 male and 4 female patients included lesions on both the upper and lower extremities, from all WHO lesion categories [1]. Clinical features of these patients are summarised in Table 1. In BU patients displaying ulcerated lesions, punch biopsies were taken 1 cm inside the outer margin of the induration surrounding the ulcer. Otherwise, punch biopsies were collected from the non-ulcerated centre of the skin lesions. After removal, tissues were fixed in 10% neutral buffered formalin, transported, embedded in paraffin and sectioned. The normal human skin tissue blocks were made from 4 mm punch biopsies collected by The Whiteley Clinic or surgical removal specimens (< 1cm^2^) collected by AMS Biotechnology-collaborated research clinical centres.

Nine serial tissue sections of each patient were (immuno-)histochemically stained in the following order: haematoxylin-eosin, fibrin, thrombomodulin (CD141), CD61, CD31 (Platelet endothelial cell adhesion molecule; PECAM-1), SMA (perivascular cell marker), Ziehl-Neelsen, TF and vWF. For immunohistochemical staining, 5-µm skin tissue sections were deparaffinised, endogenous peroxidase quenched, epitope unmasked (either in preheated pH 6 citrate buffer or treated with proteinase K, DAKO) and blocked with normal horse serum (Vector Laboratories). The tissue sections were incubated with primary antibody overnight at 4°C and biotin-conjugated horse anti-mouse/ rabbit IgG (Vector Laboratories) for 30 minutes at room temperature. Primary antibodies were as follows: CD31 (M0823, DAKO), thrombomodulin (M0617, DAKO), SMA (NCL-SMA, Novocastra), CD61 (M0753, DAKO), vWF (ab6994, Abcam), TF (TF218; generous gift from Professor Wolfram Ruf, Scripps Research Institute) and Fibrin (59D8 [96]; generous gift from Professor Charles Esmon, Oklahoma Medical Research Foundation). Staining was performed using VECTASTAIN Elite ABC kit and Vector NovaRED peroxidase substrate (Vector Laboratories). Counterstaining was performed with Shandon Harris Haematoxylin (Thermo Fisher Scientific). Anti-TF and vWF antibodies and their respective matched isotype controls were introduced to healthy skin tissue slides to rule out unspecific signals. High resolution images of all slides were scanned using either Aperio slide scanner (Leica Biosystems), Pannoramic digital slide scanners (3DHISTECH) or Hamamatsu slide photometry system (Hamamatsu Photonics). Scanned images were further analysed using ImageScope software (Leica Biosystems) or CaseViewer (3DHISTECH). In some cases, photographs were taken with Micropix microscope camera (acquisition software Cytocam) attached to a Yenway CX40 laboratory microscope (Micropix).

### Vessel identification and marker analysis

Regions of BU punch biopsy samples containing as many non-necrotic blood vessels as possible were defined by a pathologist prior to the start of analysis. This decision was made based on the number of blood vessels present that were positive for SMA or CD31 and whether there was any submacroscopic appearance of coagulative necrosis present in the corresponding H&E section. Two patient biopsies showed two distinct regions that contained plentiful non-necrotic blood vessels, therefore a total of ten representative areas were analysed for each biomarker. Once the regions were chosen, all blood vessels within them were analysed, regardless of whether nearby tissue displayed local signs of necrosis, in order to facilitate an unbiased analysis. Throughout this manuscript, we refer to these as “pre-defined” regions. Vessels within these pre-defined regions were initially identified by morphology and positive staining for SMA using the region of interest (ROI) tool on NIS Elements Basic Research (Nikon, Tokyo, Japan) (version 4.6). This allowed for the same vessels to be easily identified in the other sections. Any structure that had vessel morphology and that stained positively for the known endothelial markers CD31, thrombomodulin, or vWF in their corresponding sections were also identified and then tracked in the corresponding sections. Sweat glands stain for SMA, but do not have endothelium and do not stain for CD31, thromobomodulin or vWF [97, 98], therefore the morphology of singly SMA^+^ structures were carefully considered before assigning them as vessels. Fibrin deposition and TF expression pattern around these vessels, if identifiable, was categorised. Note that not every vessel could be identified within all sections, therefore the number of vessels analysed always represents the number that could be identified through all the relevant sections, and therefore varies from analysis to analysis.

To analyse fibrin deposition that was largely seen within the tissues, two different approaches were taken. First, the distance that fibrin staining extended from the basement membrane was categorised based on measurements taken with Aperio ImageScope (version 12.3.3). Vessels with no fibrin staining within 20 μm were scored as 0. Fibrin-positive vessels received increasing scores the further the staining extended (1; <20 μm radius, 2; 20-30 μm radius, 3; >30 μm radius). Second, fibrin staining around each vessel was quantified using the calibrated ruler tool of NIS Elements Basic Research (version 4.6 and 5.21.01). Here, the threshold for positive staining was first defined within random images, which was then applied to all vessels. Then, a 20 μm area around all vessels was measured and traced again with the ROI tool. The area of each original vessel was subtracted from the area of its 20 μm radius in order to give the area in which fibrin staining was to be quantified. Any staining within this was then calculated as a percentage of the 20 μm area around each vessel.

To analyse the unusual TF staining pattern observed, TF staining intensity within 20 μm was quantified as above for fibrin using the calibrated ruler tool of NIS Elements Basic Research (version 5.21.01). In addition, the distance that the TF signal extended from the individual vessel’s basement membrane was measured by CaseViewer (version 2.2).

### Reagents

Synthetic mycolactone A/B (generous gift of Professor Yoshito Kishi, Harvard University) [99] and its solvent control dimethyl sulfoxide (DMSO, Sigma) were used in cell-based studies. Human recombinant IL-1β was from Gibco.

### Cell culture and treatment

Primary human dermal microvascular endothelial cells (HDMEC; LONZA), human umbilical vein endothelial cells (HUVECs, PromoCell) and human dermal lymphatic endothelial cells (HDLEC; PromoCell) from two donors were used and cultured according to manufacturer’s recommendation in Endothelial cell growth medium MV2 (PromoCell). Subconfluent cells were treated with either 10 ng/mL mycolactone or DMSO equivalent to mycolactone dose for 24 hrs or as indicated in figure legend.

### Vascular permeability assay

Endothelial cells were seeded on cell culture inserts containing 0.4 (Millipore) or 1 µm pores (Falcon) with a polyethylene terephthalate membrane. Cells were treated as indicated, into both the insert and receiver wells. After 24 hrs, fluorescein isothiocyanate (FITC)-conjugated dextran (70 kDa, Millipore) was applied to each insert for 20 minutes. The fluorescence intensity of the solution in the lower chambers was then assessed by a fluorescent plate reader (FLUOstar Omega, BMG Labtech) with excitation/ emission wavelength at 485/ 530 nm. Fluorescence intensity was normalised to untreated control wells with an intact monolayer of endothelial cells (minimum) and expressed as a percentage of subtracted value obtained from wells where the insert had no cells (maximum).

### Cell viability assay

Endothelial cells (4 x 10^3^ cells) were seeded onto 96-well plates and treated with a variety of doses of mycolactone the following day. Serial dilutions of mycolactone from 50 to 0.098 ng/mL or solvent control equivalent to highest mycolactone dose (0.1% DMSO) was applied to the cells; on the fifth day viability was assayed using alamarBlue Cell Viability Reagent (Invitrogen) with excitation/ emission wavelength at 560/ 590 nm by a plate reader (FLUOstar Omega, BMG Labtech). Values were normalised to the control wells treated with DMSO and are presented as survival rate (%).

### Immunochemical analysis

Western blot analysis was carried out using standard techniques, following cell lysis with 1X RIPA buffer (Sigma) supplied with proteinase inhibitor cocktail (Sigma), separation by electrophoresis on 10% polyacrylamide gels and transfer to PVDF membranes (ThermoFisher Scientific). Antibodies used were TM (sc-13164, Santa Cruz), TFPI (AF2974, R&D Systems) and GAPDH (AM4300, Ambion). Immunofluorescence staining was performed on paraformaldehyde-fixed cells after permeabilising with 0.5% Triton X-100 for 2 minutes. Antibodies used were vWF (ab6994, abcam) and Alexa Fluor 488 goat anti-rabbit IgG (H+L) (ThermoFisher Scientific). Nuclei were visualised with DAPI. Images were acquired with Nikon A1M confocal laser scanning unit attached to an Eclipse Ti-E microscope.

To assess the TFPI protein level in conditioned medium, an in-house ELISA was performed. In brief, samples or TFPI recombinant protein standards (ranging from 1000 to 8 ng/mL, 2974-PI. R&D Systems) were coated onto immunoplates (MaxiSorp, Nunc). Following overnight incubation, individual wells were blocked with 1% bovine serum albumin. Antibodies used were TFPI (AF2974, R&D Systems), HRP Rabbit anti-goat IgG (H+L), and Avidin anti-HRP (Invitrogen). The reaction was developed with 3, 3’, 5, 5’-tetramethylbenzidine substrate (Invitrogen), stopped by 1M H_2_SO_4_ and read at 450 nm by a plate reader (FLUOstar Omega, BMG Labtech).

To determine the effect of mycolactone on the release of vWF from Weibel-Palade bodies, HDMECs were pre-incubated with DMSO or 7.8ng/mL mycolactone for 24 hours, washed twice with serum free medium, then stimulated with 2 U/mL thrombin for 10 minutes. vWF protein levels were then quantified using an in-house vWF ELISA. Anti-human vWF antibody (A0082, Dako) was coated onto immunoplates overnight at 4°C and blocked with 2% BSA in PBS for 1 hour at room temperature. After blocking, the plate was washed three times with wash buffer (0.05% (v/v) Tween-20 in PBS), samples and standards (NIBSC) ranging from 1000 mIU/mL - 4 mIU/mL were added to the wells and the plate was incubated at room temperature for 2 hours. Then, vWF was detected with HRP-conjugated rabbit anti-human vWF (P0226, Dako) followed by 3, 3’, 5, 5’-tetramethylbenzidine substrate (Invitrogen), stopped by 1M H_2_SO_4_ and read at 450 nm by a plate reader (FLUOstar Omega, BMG Labtech).

### Statistical analysis

Statistical analysis was carried out using GraphPad Prism version 8 (San Diego, USA). Categorical data of analysed vessels was accessed using Chi-square test of association. Yate’s correction for continuity was introduced to TF categorical data set as some observed values were below 5. Fibrin intensity per TM^+^ or TM^−^ vessel was assessed using Mann Whitney *U* non-parametric test. Fibrin intensity versus TF intensity or distance to BM per vessel with TF seen outside of BM was assessed using D’Agostino-Pearson normality test; as data set displayed a non-Gaussian distribution, correlation used the method of Spearman. Data otherwise was accessed using a one-way ANOVA and Dunnett’s multi-comparison test. Unless otherwise indicated, statistical comparison for *in vitro* assays was vs DMSO-treated controls.

## Data Availability

All data are fully available without restriction. All relevant data are within the manuscript and its Supporting Information files

## Acknowledgments

We would like to thank Lucia Lozano White (University of Surrey) for preparing tissue sections from healthy human skin tissue blocks, Dr Yoshito Kishi (Harvard University, USA) for the gift of synthetic mycolactone A/B, Prof Wolfram Ruf (Scripps Research Institute) and Prof Charles Esmon (Oklahoma Medical Research Foundation) for antibodies against TF and thrombomodulin, respectively. We thank Dr Belinda Hall and Dr Jane Newcombe for critical reading of the manuscript and Prof Mark Wansbrough-Jones (St Georges University of London) for helpful discussions leading to the testing of lymphatic endothelial cells.

## Supporting information Captions

**S1 Fig.**
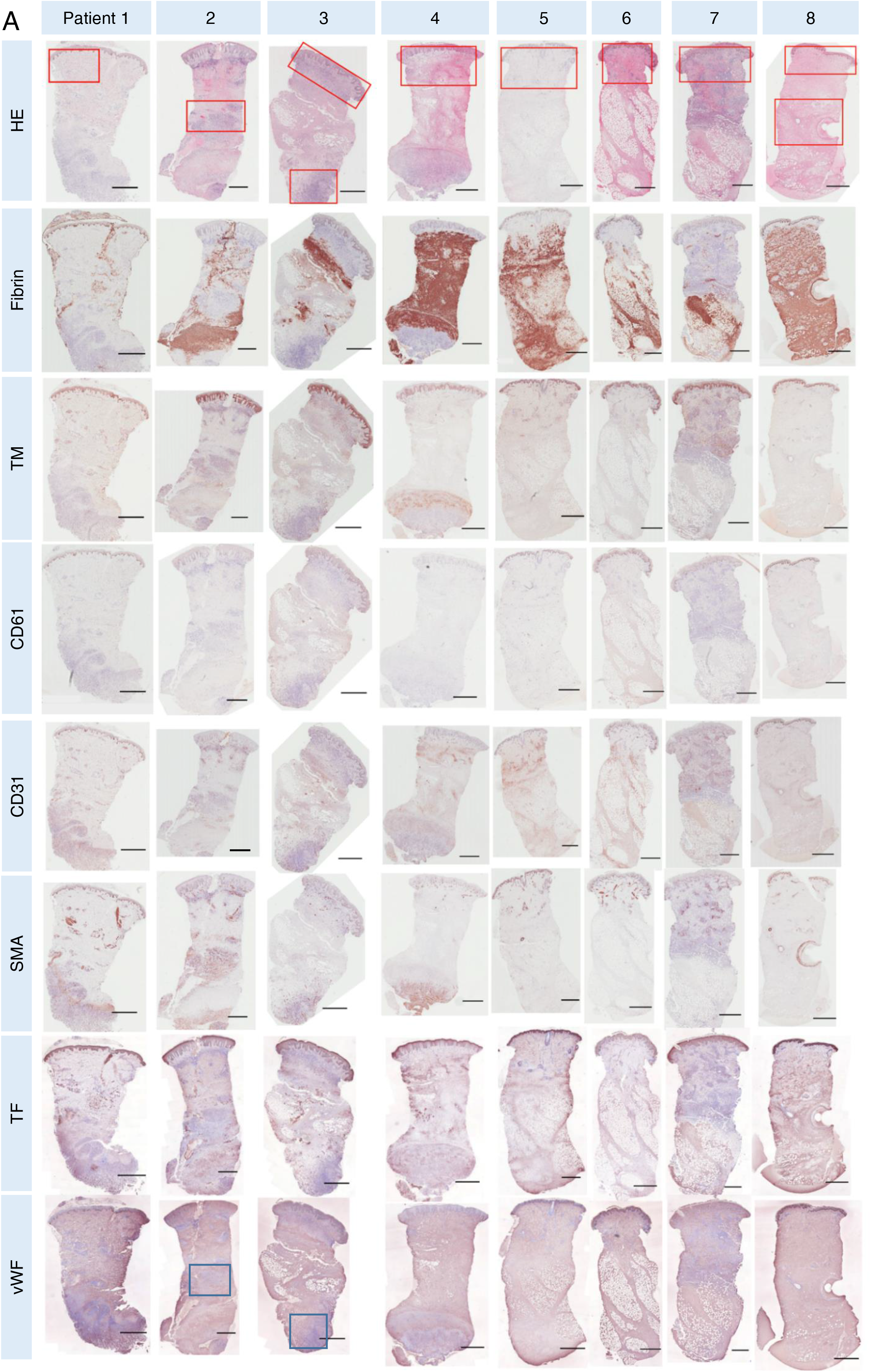
The expression patterns of haemostatic modulators in 8 BU patient skin biopsies. Histological sections from 8 BU patient punch biopsies stained with eosin (H&E) or antibody against fibrin, TM, CD61, CD31, SMA, TF and vWF and counterstained with Haematoxylin. The defined regions identified by a pathologist are outlined in red; vessels in these areas were tracked and analysed for this study. Immune cell infiltration regions containing vWF seen in micro vessel lumen outlined in blue. Scale bar = 1 mm.

**S2 Fig.**
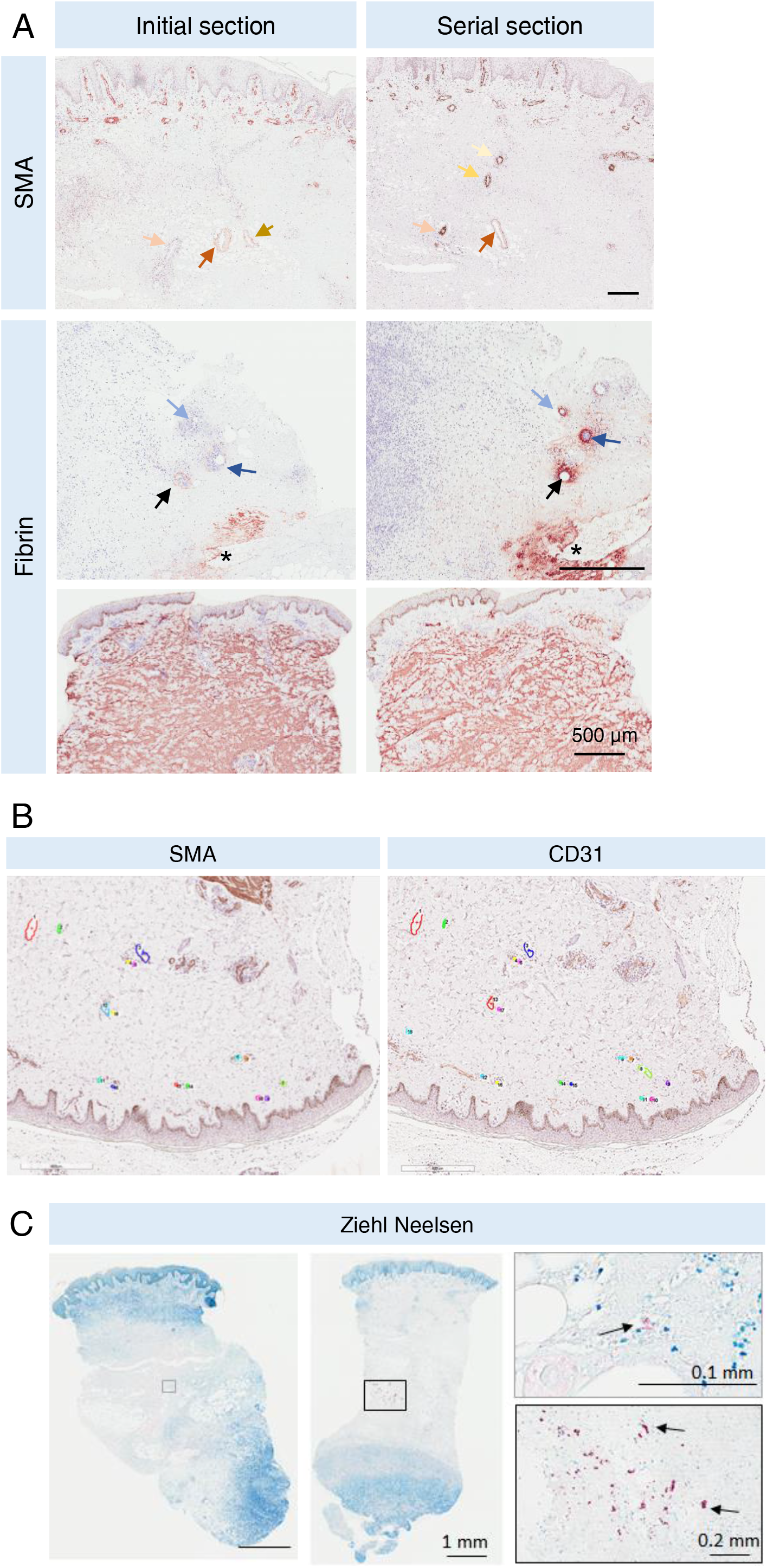
General features of BU patient skin biopsies. **A.** Comparison of the staining patterns with the same anti-SMA or anti-fibrin antibody and conditions seen for the same punch biopsies at different positions in the tissue block. The “initial section” displays those performed for Ogbechi et al., 2015 [28], whereas the “serial section” was performed for the present work. Arrows in different colours label the same vessel identified in different tissue sections. Scale bar = 500 µm. Note how the vessel phenotype can vary even over small distances. **B.** An example of vessel identification and labelling (in colours with individual number indicated) in serial tissue sections stained with anti-SMA and CD31 antibody. **C.** Mycobacterial clusters (in purple, indicated with arrows) are identified in histological sections from 2 BU patient punch biopsies with Ziehl-Neelsen staining. Scar bars as indicated.

**S3 Fig.**
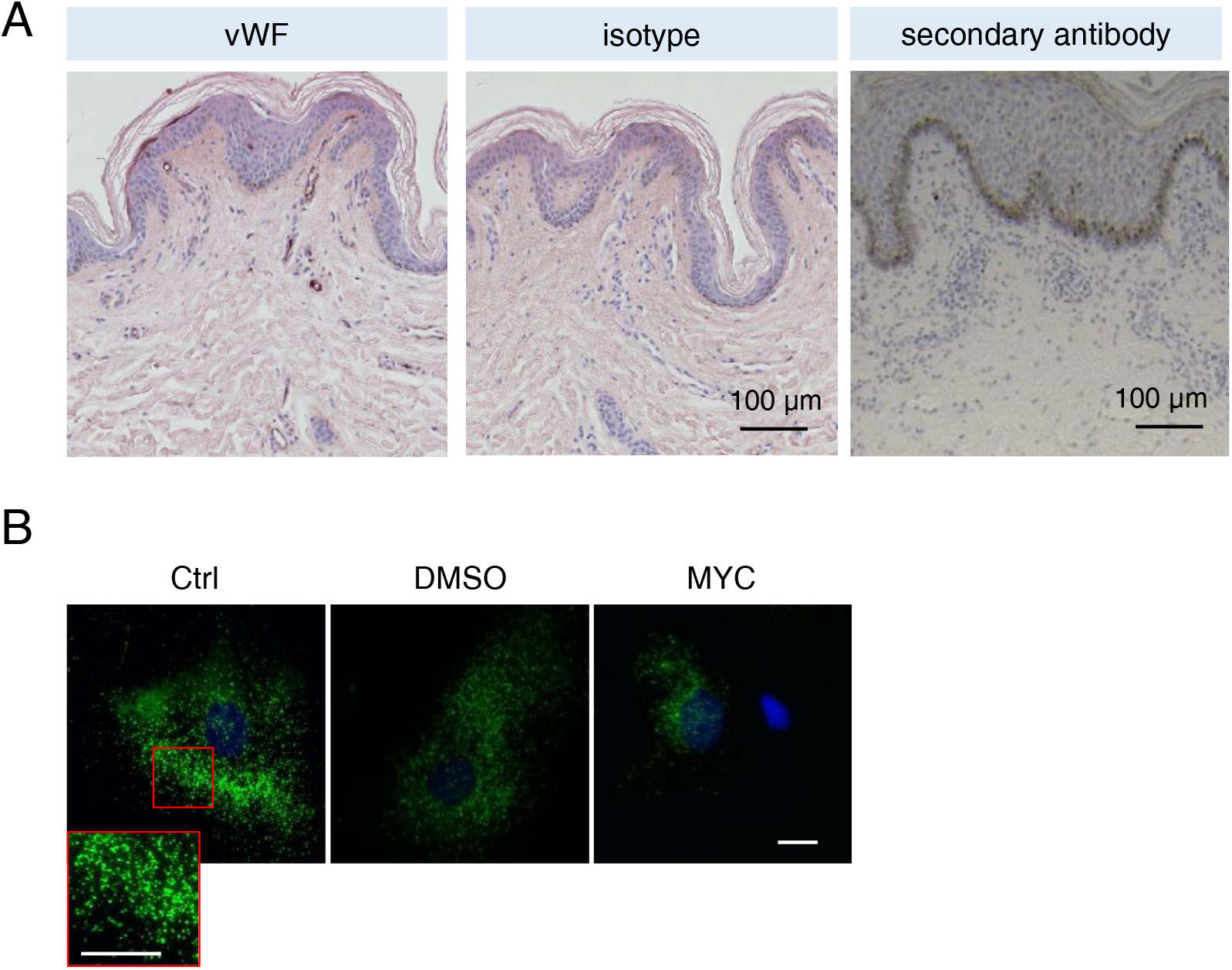
vWF expression in BU patient specimen and in primary HDMECs. **A.** Histological sections of a healthy subject or BU patient stained with anti-vWF antibody, the respective isotype control or secondary antibody alone and counterstained with Haematoxylin. **B.** HDMECs were treated with 10 ng/mL of mycolactone (MYC), 0.02% DMSO or untreated (Ctrl) for 24 hours. Cells were fixed, permeabilised and immunostained with anti-vWF antibody. vWF-containing granules are shown in green and nuclei stained with DAPI (blue). Scale bar = 20 µm.

**S4 Fig.**
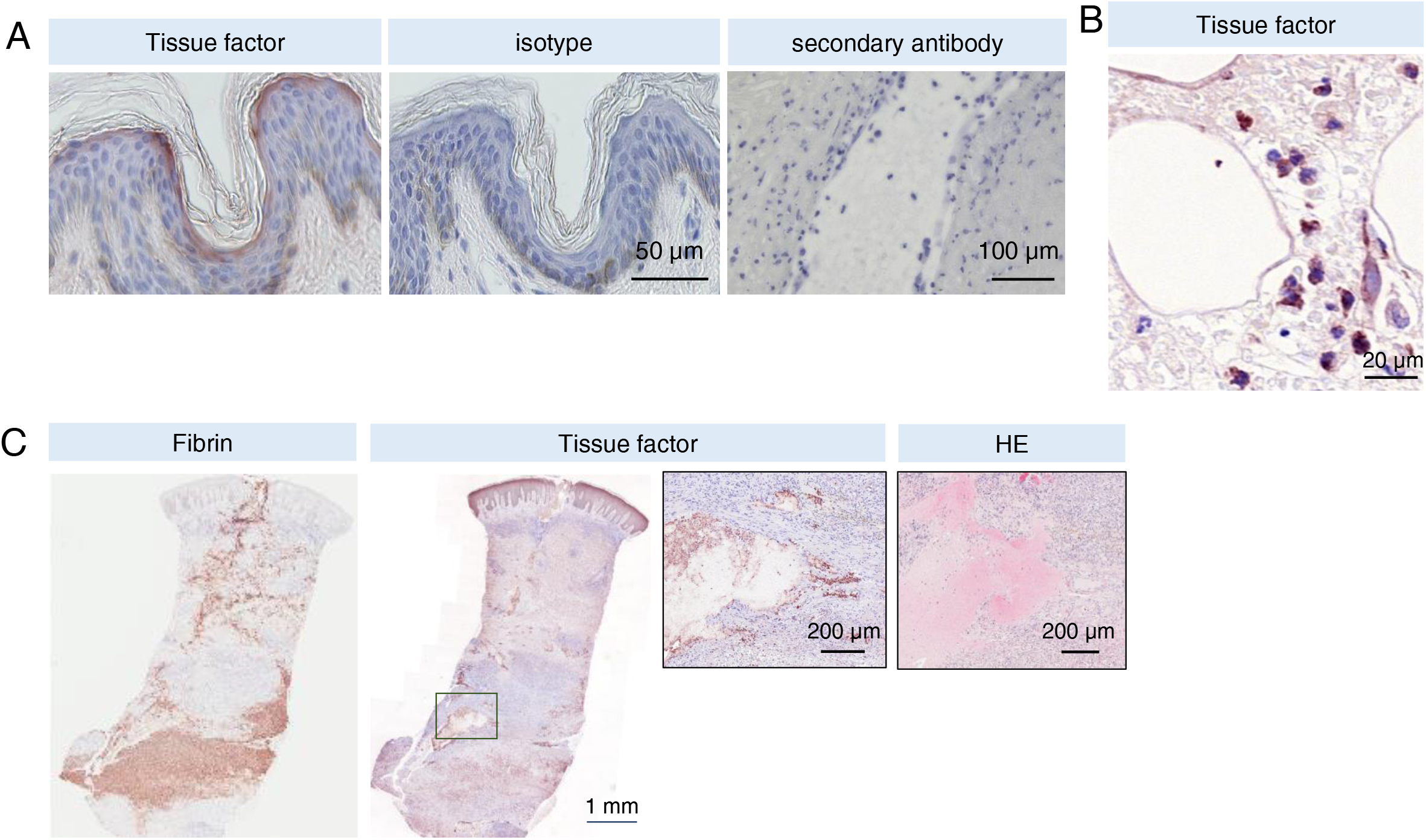
Tissue factor (TF) expression patterns in BU patient specimen. **A.** Histological sections of a healthy subject or BU patient stained with anti-TF antibody, the respective isotype control or secondary antibody alone and counterstained with Haematoxylin. **B & C.** Histological sections stained with Haematoxylin and eosin or antibodies against TF and fibrin and counterstained with Haematoxylin in BU patients. TF-positive activated leukocytes in necrotic subcutis (**B**) and structures at the edge of the haemorrhagic area (**C**). Scale bar as indicated.

**S5 Fig.**
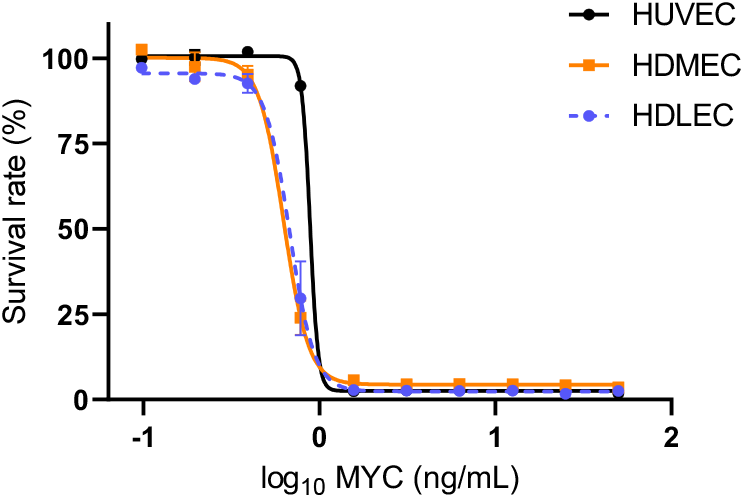
Susceptibility of endothelial cells to mycolactone. Cell viability of HDLECs, HDMECs and HUVECs exposed to a variety doses of mycolactone for 5 days was determined using alamarBlue assay and presented as a % where 100% is the value obtained from cells treated with solvent control DMSO.

